# Association of pre-existing maternal cardiovascular diseases with neurodevelopmental disorders in offspring: a cohort study in Sweden and British Columbia, Canada

**DOI:** 10.1101/2023.01.24.23284797

**Authors:** M. Zakir Hossin, Lorena Fernández de la Cruz, Kyla A McKay, Tim Toberlander, Anna Sandström, Neda Razaz

## Abstract

**Importance:** Maternal cardiac disease is associated with impaired placentation and adverse neonatal outcomes, but whether it increases the risk of subsequent neurodevelopmental disorders in offspring remains unknown.

**Objective:** To assess the associations between pre-existing maternal cardiovascular disease (CVD) and attention-deficit/hyperactivity disorder (ADHD), autism spectrum disorder (ASD), and intellectual disability (ID) in offspring.

**Design:** Population-based cohort study.

**Setting:** Sweden and British Columbia (BC), Canada.

**Participants:** Singletons live-born without major malformations between 1990 and 2019. The Swedish Medical Birth Register and the BC Vital Statistics were linked to hospital and physician billing records for identification of exposure and outcomes. All children were followed from age 1 until the outcome, death, emigration, or December 2020.

**Exposure:** A composite of pre-conception maternal CVDs: cerebrovascular disease, arrhythmia, heart failure, valvular, and congenital heart diseases.

**Main Outcomes and Measures:** Diagnoses of ADHD, ASD, and ID. The incidence of ADHD, ASD, and ID, comparing offspring of mothers with versus without CVD, was calculated as adjusted hazard ratios (aHRs). These results were compared to models using paternal CVD as negative-control exposure. Fixed-effect meta-analysis was performed to combine the aHRs from Sweden and BC.

**Results:** We analysed 3,587,257 offspring (48.9% female) – 2,699,675 in Sweden and 887,582 in BC. The meta-analysis suggests that compared with offspring of mothers without CVD, offspring of mothers with CVD had 1.15-fold higher aHRs of ADHD (95% CI: 1.10-1.20) and ASD (95% CI: 1.07-1.22). No association was found between maternal CVD and ID. Stratification by maternal CVD subtypes showed increased hazards of ADHD for maternal heart failure (aHR 1.31; 95% CI: 1.02-1.61), cerebrovascular disease (aHR 1.20; 95% CI: 1.08-1.32), congenital heart disease (aHR 1.18; 95% CI: 1.08-1.27), arrhythmia (aHR 1.13; 95% CI: 1.08-1.19), and valvular heart disease (aHR 1.12; 95% CI: 1.00-1.24). Increased hazards of ASD were observed for maternal cerebrovascular disease (aHR 1.25; 95% CI: 1.04-1.46), congenital heart disease (aHR 1.17; 95% CI: 1.01-1.33), and arrythmia (HR 1.12; 95% CI: 1.01-1.21). Paternal CVDs, except for cerebrovascular disease, did not show associations.

**Conclusion and Relevance:** Pre-existing maternal CVD was associated with excess risk of ADHD and ASD in offspring. The associations were broadly robust to genetic confounding, warranting further research to identify the underlying intrauterine mechanisms.

## Introduction

Cardiovascular disease (CVD) is a major cause of pregnancy complications, morbidity, and mortality in pregnant women^1^. The prevalence of CVD among women of childbearing age is rising globally^2,3^ due to the growing prevalence of classic cardiovascular risk factors, the increased survival of women with congenital and other heart diseases, and the rising trend to delay motherhood to later ages of reproduction^3^. The high metabolic demands of the mother and the fetus during pregnancy require substantial physiologic and hemodynamic adaptations^4^ which might not be well-tolerated in pregnancies with a pre-established cardiovascular history. Pathophysiologically, maternal CVD may alter the placental function and hamper utero-placental delivery of oxygen and nutrients to the fetus, thereby adversely impacting fetus’ composition and growth, including brain development^5,6^.

A cumulative body of epidemiological literature suggests that heart disease in pregnancy is associated with adverse maternal and neonatal health outcomes^7,8^, but the longer-term effects of chronic maternal CVDs on the developing offspring, especially on cognitive and neurodevelopmental outcomes, remain to be explored^9^. While neurodevelopmental disorders (NDDs), such as attention-deficit/hyperactivity disorder (ADHD)^10^, autism spectrum disorder (ASD)^11^, and intellectual disability (ID), are highly heritable phenotypes^12^, a variety of environmental factors may also contribute to their aetiology^13^ and could be the target of prevention or early intervention strategies.

Previous studies have documented elevated risks of NDDs in offspring of mothers with pre-pregnancy obesity^14–17^, pre-gestational and gestational diabetes^15,18^, and hypertensive disorders during pregnancy^19,20^. To our knowledge, no investigation has explored the association between pre-pregnancy maternal CVD and long-term NDD outcomes in offspring. In this population-based study of nearly 3.6 million children born in Sweden or British Columbia (BC), Canada, we examined the association of maternal pre-existing CVDs with ADHD, ASD, and ID in offspring. In addition, we employed a family-based design using paternal CVDs as negative controls to explore whether a possible association might be confounded by unmeasured genetic factors.

## Methods

### Data Sources

The data were primarily derived from the Swedish Medical Birth Register^21^ and the British Columbia Vital Statistics Birth file^22^ that cover more than 98% of all births occurring in Sweden and BC, respectively. The Swedish Medical Birth Register retrieves information on pre-pregancy, perinatal, and neonatal characteristics based on medical records from antenatal and delivery health care. In Sweden, we cross-linked the Medical Birth Register to several national registers: the National Patient Register (inpatient and outpatient specialist care)^23,24^, the Cause of Death Register^25^, the Prescribed Drug Register^26^, the Total Population Register^27^, and the Education Register^28^. In BC, the British Columbia Vital Statistics Birth database^22^ was linked to: the Discharge Abstract Database (hospitalizations)^29^, the Medical Services Plan physician billing data (outpatient physician visits)^30^, the PharmaNet (drug exposure)^31^, and the Central Demographics File^32^ (see eTable 2 for details of all registries). The study was approved by the Ethics Review Authority in Sweden and the Ethics Committee at the University of British Columbia, Canada.

### Study Population

We identified 3,074,630 live singleton births with ≥ 22 completed gestational weeks from the Swedish Medical Birth register from 1 January 1990 to 31 December 2019 and 1,176,936 live singleton births at ≥ 22 completed gestational weeks from the BC Vital Statistics Births from 1 January 1992 to 31 December 2019. We excluded infants with any major malformations (International Classification of Disease [ICD]-9 codes: 740–759; ICD-10 codes: Q00–Q99), births where either the mothers or the children had missing national registration number, and children who had died, emigrated or been diagnosed with any study outcomes before reaching their 1^st^ birthday (start of follow-up). Moreover, in the unexposed group, we excluded children of mothers with diagnostic ICD-codes for any CVD that were not part of our definition of maternal CVD (see eTable 1 for codes), except those with codes for maternal hypertensive diseases. Of the children who met the eligibility criteria for inclusion, 2,699,675 (92%) in Sweden and 827,896 (94%) in BC had complete data on the study variables and were subject to statistical analyses. For comparison with maternal CVD, we also linked 2,672,229 (91%) children in Sweden and 811,481 (91%) children in BC to their biological fathers with available information on CVD diagnoses (eFigure 1).

### Exposure

The primary exposure was maternal CVD prior to conception (i.e., date of birth minus gestational age), including primary or secondary diagnoses of: cerebrovascular disease, arrhythmia, heart failure, valvular heart disease, and congenital heart disease (eTable 1). A composite maternal CVD exposure included any of these conditions. We further examined each subtype separately. In Sweden, maternal CVDs were identified as one or more ICD-9 or ICD-10 codes for CVD in the National Patient Register, including inpatient (since 1987) and outpatient specialist care (since 2001). In BC, these were identified as one or more ICD codes for CVD in the Discharge Abstract Database or two or more records for CVD in the Medical Services Plan data between 1985 and 2019. Paternal CVD was defined using the same ICD codes as for mothers. While the diagnoses in Sweden only include specialist care data, those in BC include both specialist and primary care. The ICD-10 codes became available in Sweden in 1997 and in BC from 2001, prior to which the ICD-9 codes were used for disease identification. The validity of cardiovascular diagnoses in the Swedish inpatient register has been shown to be generally high^24^.

### Outcomes

Offspring diagnoses of ADHD, ASD, and ID were extracted from the 1^st^ birthday until 31 December, 2020 from the National Patient Register in Sweden and the Discharge Abstract Database and the Medical Services Plan in BC (eTable 2). ADHD cases were further identified through records of dispensation of ADHD medication, using the Anatomical Therapeutic Chemical (ATC) codes retrieved from the Prescribed Drug Register in Sweden (since July 2005) ^26^ and PharmaNet in BC (since 1996; eTable 2)^31^.

### Covariates

The covariates selected a priori in the study were infant characteristics including sex, birth year, preterm birth (<37 gestational weeks), and small-for-gestational age (<10^th^ percentile of the standardized birth weight distribution), as well as maternal characteristics including mother’s age at delivery, parity, region of birth, marital status/cohabitation with partner, educational level (Sweden only), smoking during early pregnancy (Sweden only; self-reported at first prenatal visit or at 30 to 32 gestational weeks), pre-gestational diabetes, pre-gestational hypertension, and both parents’ history of any neurodevelopmental or psychiatric disorders. Preterm birth and small-for-gestational age were considered as potential mediators, as they were previously shown to be associated with maternal heart disease^7^ and child NDDs^13^. The registers from which information on specific covariates was obtained are shown in eTable 2.

### Statistical Analyses

We assessed the distribution of neonatal and maternal characteristics by maternal CVD status. Cox proportional hazard regression models were used to estimate the incidence rates and hazard ratios (HRs) with 95% confidence intervals (CIs) by overall maternal CVD and its subtypes. For each NDD outcome, follow-up commenced at age 1 and continued until the diagnosis of the outcome, death, emigration or 31 December, 2020, whichever came first. Child’s attained age was used as the underlying time metric in all Cox models. We controlled for child’s age, sex, and birth year in a minimally adjusted model, followed by a fully adjusted model additionally controlling for maternal characteristics including mother’s age at delivery, parity, education, region of birth, marital status/cohabitation with partner, smoking during early pregnancy, pre-gestational diabetes, pre-gestational hypertension, and parental history of any neurodevelopmental or psychiatric disorders. Since children of the same mother comprise a cluster, possible intra-cluster correlations were accounted for by estimating robust standard errors.

We also performed a negative control analysis^33^, using paternal CVD status as exposure, to detect if the estimated associations between maternal CVDs and offspring outcomes were confounded by unmeasured common causes, including genetics (eFigure 2). The paternal comparison models were adjusted analogously to models evaluating maternal CVD, with additional adjustment for maternal CVD.

In addition, we undertook a counterfactual mediation analysis with preterm birth as a mediator. We used Generalized Linear Poisson regression models to decompose the total effect of maternal CVD on ADHD, ASD or ID into natural indirect effect (i.e., the effect that goes through preterm birth) and natural direct effect (i.e., the effect that goes through other mechanisms). Small-for-gestational age and ID were weakly associated with the exposure in our initial investigation and were excluded from the final mediation analysis.

All analyses were performed separately for Sweden and BC, with the fully adjusted HRs subsequently combined through fixed-effect meta-analysis using the inverse variance method. The BC data was analysed in SAS version 9.4. Stata version 17.0 was used to analyse the Swedish data and perform the meta-analysis.

### Sensitivity Analyses

We perfomed several sensitivity analyses. First, information on maternal weight and height was recorded in the Swedish Medical Birth Register from 1992 and was missing in 18% of women, therefore statistical adjustment for maternal body mass index (kg/m^2^; BMI) was only made in an additional analysis. Second, to investigate if the associations were robust to diagnostic changes of the exposure and outcomes over time, we repeated the main analysis by restricting the Swedish sample to children born from 1997 and the BC sample to children born from 2001, when the the ICD-10 codes became available. Third, aggregated data on average neighbourhood income was available in BC (missing 2.7%), therefore, we used this information by adjusting for parental neighbourhood income quintiles in maternal-offspring associations. Fourth, considering the co-occurrence of ASD and ID as reported in previous literature^34^, we further examined if maternal CVD was associated with offspring ASD without ID. Fifth, for the paternal-offspring associations, we treated paternal age as an additional confounder in a sensitivity analysis, as advanced paternal age might be a common cause of paternal CVD and offspring NDD^35^.

### Missing Data

Children with missing data on any study variables were excluded (Sweden 8.2%; BC 6.2%) from the main analysis. In a supplementary analysis of the Swedish data, we created 10 imputed datasets under the assumption of missing at random, using the multiple imputation by chained equations procedure^36^. The estimates from the imputed datasets were combined by Rubin’s rule.

## Results

Of the 2,699,675 children analysed in Sweden (48.9% female), a total of 22,775 (0.8%) were born to mothers with pre-existing CVD. Among women with CVD, 13,064 (57%) were diagnosed with arrhythmia, 5534 (24%) with congenital heart disease, 3414 (15%) with cerebrovascular disease, 2069 (9%) with valvular heart disease, and 515 (2%) with heart failure. In BC, we analysed 887,582 children (49.1% female) of which 22,010 (2.5%) were born to mothers with any pre-existing CVD: 14,346 arrhythmia (65%), 4168 congenital heart disease (19%), 2111 cerebrovascular disease (10%), 2712 valvular heart disease (12%), and 718 heart failure (3%).

Table 1 shows the characteristics of the two study populations by maternal CVD status. In both Sweden and BC, mothers with CVD were more likely to be older, multiparous, born in Sweden/BC, and live without a partner, compared with mothers without CVD. They also had higher frequency of pre-gestational diabetes, hypertension, and history of any neurodevelopmental or psychiatric disorder. Offspring of mothers with CVD were more likely to be born preterm.

**Table 1.** Maternal and offspring characteristics according to maternal pre-existing cardiovascular diseases: singleton offspring live-born without major malformations in Sweden 1990 to 2019 and in British Columbia, Canada 1992 to 2019

| Characteristics | Sweden |  |  | British Columbia, Canada |  |  |
| --- | --- | --- | --- | --- | --- | --- |
|  | Total<br>(N=2 699 675) | Maternal CVD |  | Total<br>(N=887 582) | Maternal CVD |  |
|  |  | No<br>(N=2 676 900) | Yes<br>(N=22 775) |  | No<br>(N=865 572) | Yes<br>(N=22 010) |
| Offspring characteristics % (N) |  |  |  |  |  |  |
| Sex |  |  |  |  |  |  |
| Male | 51.2 (1 380 754) | 51.1 (1 369 074) | 51.3 (11 680) | 50.9 (451 766) | 50.9 (440 502) | 51.2 (11 264) |
| Female | 48.8 (1 318 921) | 48.9 (1 307 826) | 48.7 (11 095) | 49.1 (435, 816) | 49.1 (425 070) | 48.8 (10 746) |
| Birth year |  |  |  |  |  |  |
| <1995 | 18.1 (489 118) | 18.2 (488 405) | 3.1 (713) | 12.2 (108 741) | 12.5 (107 788) | 4.3 (953) |
| 1995-1999 | 14.6 (395 250) | 14.7 (394 033) | 5.3 (1217) | 18.0 (159 311) | 18.1 (156 830) | 11.3 (2481) |
| 2000-2004 | 14.9 (403061) | 15.0 (400 913) | 9.4 (2148) | 16.8 (148 859) | 16.8 (145 651) | 14.6 (3208) |
| 2005-2009 | 16.5 (446 162) | 16.5 (441 995) | 18.3 (4167) | 17.6 (155 860) | 17.5 (151 776) | 18.6 (4084) |
| 2010-2014 | 17.9 (483 033) | 17.8 (476 523) | 28.6 (6510) | 17.8 (158 357) | 17.7 (153 264) | 23.1 (5093) |
| 2015-2019 | 17.9 (483051) | 17.8 (475 031) | 35.2 (8020) | 17.6 (156 454) | 17.4 (150 263) | 28.1 (6191) |
| Preterm birth |  |  |  |  |  |  |
| No (37-44 weeks) | 95.7 (2 582 605) | 95.7 (2 561 143) | 94.2 (21462) | 95.1 (844 077) | 95.1 (823 450) | 93.7 (20 627) |
| Yes (<37 weeks) | 4.3 (117 070) | 4.3 (115 757) | 5.8 (1313) | 4.9 (43 505) | 4.9 (42 122) | 6.3 (1383) |
| Small-for-gestational age |  |  |  |  |  |  |
| No | 93.7 (2 530 487) | 93.7 (2 509 193) | 93.5 (21 294) | 97.0 (860 618) | 97.0 (839 184) | 97.4 (21 434) |
| Yes | 6.3 (169 188) | 6.3 (167 707) | 6.5 (1481) | 3.0 (26 964) | 3.0 (26 388) | 2.6 (576) |
| Maternal characteristics % (N) |  |  |  |  |  |  |
| Maternal age at delivery (yrs) |  |  |  |  |  |  |
| ≤19 | 1.0 (233) | 1.6 (43 648) | 1.1 (195) | 3.6 (31 689) | 3.6 (31 129) | 2.5 (560) |
| 20-24 | 11.5 (2609) | 14.6 (390 428) | 12.3 (2177) | 14.7 (130 718) | 14.8 (128 080) | 12.0 (2638) |
| 25-29 | 29.9 (6819) | 33.1 (886 069) | 29.1 (5164) | 29.1 (258 614) | 29.2 (252 909) | 25.9 (5705) |
| 30-34 | 33.3 (7592) | 32.5 (870 408) | 33.3 (5912) | 33.0 (292 339) | 32.9 (284 778) | 34.4 (7561) |
| ≥35 | 24.3 (5522) | 18.2 (486 347) | 24.2 (4289) | 19.6 (174 222) | 19.5 (168 676) | 25.2 (5546) |
| Parity |  |  |  |  |  |  |
| 1 | 42.9 (1 159 845) | 43.0 (1 151 123) | 38.3 (8722) | 46.3 (411 422) | 46.6 (402 965) | 38.4 (8457) |
| 2 | 37.1 (1 001 045) | 37.0 (992 255) | 38.6 (8790) | 36.2 (321 296) | 36.1 (312 816) | 38.5 (8480) |
| 3 | 14.0 (377 224) | 14.0 (373 717) | 15.4 (3507) | 12.2 (108 347) | 12.1 (104 920) | 15.6 (3427) |
| ≥4 | 6.0 (161 561) | 6.0 (159 805) | 7.7 (1756) | 5.2 (46 517) | 5.2 (44 871) | 7.5 (1646) |
| Region of birth |  |  |  |  |  |  |
| Sweden | 79.8 (2 154 173) | 79.7 (2 134 569) | 86.1 (19 604) |  |  |  |
| Other Nordic | 2.0 (54 112) | 2.0 (53 858) | 1.1 (254) |  |  |  |
| Non-Nordic | 18.2 (491 390) | 18.3 (488 473) | 12.8 (2917) |  |  |  |
| Region of birth<sup>a</sup> |  |  |  |  |  |  |
| Canada |  |  |  | 68.2 (605 499) | 67.8 (587 095) | 83.6 (18 404) |
| Asia |  |  |  | 22.7 (201 376) | 23.0 (199 166) | 10.0 (2210) |
| Europe |  |  |  | 5.2 (46 077) | 5.2 (45 232) | 3.8 (845) |
| Others |  |  |  | 3.9 (34 630) | 3.9 (34 079) | 2.5 (551) |
| Cohabitation with partner |  |  |  |  |  |  |
| Yes | 94.3 (2 545 600) | 94.3 (2 524 248) | 93.7 (21 352) |  |  |  |
| No | 5.7 (154 075) | 5.7 (152 652) | 6.3 (1423) |  |  |  |
| Marital status<sup>a</sup> |  |  |  |  |  |  |
| Married |  |  |  | 73.2 (649 457) | 73.2 (634 042) | 70.0 (15 415) |
| Divorced/separated/widowed |  |  |  | 19.2 (170 526) | 19.2 (165 922) | 20.9 (4604) |
| Never |  |  |  | 3.3 (29 316) | 3.3 (28 392) | 4.2 (924) |
| Other |  |  |  | 4.3 (38 283) | 4.3 (37 216) | 4.9 (1067) |
| Level of education (years)<sup>b</sup> |  |  |  |  |  |  |
| ≤9 | 8.7 (233 481) | 8.6 (231 537) | 8.5 (1944) |  |  |  |
| 10-11 | 16.9 (456953) | 17.0 (454 275) | 11.8 (2678) |  |  |  |
| 12 | 24.1 (651 634) | 24.1 (645 408) | 27.3 (6226) |  |  |  |
| 13-14 | 15.1 (407 096) | 15.1 (404 048) | 13.4 (3048) |  |  |  |
| ≥15 | 35.2 (950 511) | 35.2 (941 632) | 39.0 (8879) |  |  |  |
| Smoking in early pregnancy<sup>b</sup> |  |  |  |  |  |  |
| No | 88.7 (2 394 604) | 88.7 (2 373 861) | 91.1 (20 743) |  |  |  |
| Yes | 11.3 (305 071) | 11.3 (303 039) | 8.9 (2032) |  |  |  |
| Any neurodevelopmental or psychiatric disorders |  |  |  |  |  |  |
| None | 92.9 (2 507 947) | 93.0 (2 489 533) | 80.9 (18 414) | 91.2 (809 580) | 91.5 (792 203) | 79.0 (17 376) |
| Neurodevelopmental disorders | 0.6 ( 16 465) | 0.6 (15 960) | 2.2 (505) | 0.3 (3010) | 0.3 (2796) | 1.0 (214) |
| Psychiatric disorders | 6.5 (175 251) | 6.4 ( 171 398) | 16.9 (3853) | 8.5 (74 992) | 8.2 (70 573) | 20.0 (4420) |
| Pre-gestational diabetes |  |  |  |  |  |  |
| No | 99.6 (2 688 552) | 99.6 (2 665 957) | 99.2 (22 595) | 95.1 (844 483) | 95.2 (824 449) | 91.0 (20 034) |
| Yes | 0.4 (11 123) | 0.4 (10 943) | 0.8 (180) | 4.9 (43 099) | 4.8 (41 123) | 9.0 (1976) |
| Pre-gestational hypertension |  |  |  |  |  |  |
| No | 99.3 (2 681 809) | 99.4 (2 659 422) | 98.3 (22 387) | 99.5 (883 120) | 99.5 (861 254) | 99.3 (21 866) |
| Yes | 0.7 (17866) | 0.6 (17 478) | 1.7 (388) | 0.5 (4462) | 0.5 (4318) | 0.7 (144) |

CVD, cardiovascular disease.
<sup>a</sup>Mother's region of birth and marital status were categorized differently in British Columbia, Canada.
<sup>b</sup>Information on maternal education and smoking was not available in British Columbia, Canada.

During the observation period from 1991 to 2020 in Sweden (median age at follow-up: 13-15 years), 154,399 children were diagnosed with ADHD (rate 4.1/1000 child-years), 64,436 with ASD (rate 1.7/1000 child-years), and 22,758 were diagnosed with ID (rate 0.6/1000 child-years). In BC, the corresponding number of children with ADHD, ASD, and ID during 1992-2020 (median age at follow-up: 12-13 years) were 78,333 (rate 7.1/1000 child-years), 19,018 (rate 1.6/1000 child-years), and 4132 (rate 0.4/1000 child-years), respectively. Offspring exposed to maternal CVD generally had higher rates of ADHD, ASD, and ID compared with offspring without maternal CVD. The minimally adjusted models showed that maternal CVD was associated with increased hazards of ADHD and ASD in both Sweden and BC. Paternal CVD also showed increased hazards of ADHD and ASD in Sweden and ASD in BC (Table 2).

**Table 2.** Incidence rates and hazard ratios of neurodevelopmental disorders according to pre-existing maternal and paternal cardiovascular diseases: singleton offspring live-born without major malformations in Sweden 1990 to 2019 and in British Columbia, Canada 1992 to 2019

| Sweden |  |  |  |  |  |  |  |  |  |
| --- | --- | --- | --- | --- | --- | --- | --- | --- | --- |
|  | ADHD |  |  | ASD |  |  | ID |  |  |
|  | No. of events | Rates<sup>a</sup> | Minimally adjusted HR<sup>b</sup> (95% CI) | No. of events | Rates<sup>a</sup> | Minimally adjusted HR<sup>b</sup> (95% CI) | No. of events | Rates<sup>a</sup> | Minimally adjusted HR<sup>b</sup> (95% CI) |
| **Maternal CVD (N=2 699 675)** |  |  |  |  |  |  |  |  |  |
| Composite maternal CVD |  |  |  |  |  |  |  |  |  |
| No | 153 355 | 4.1 | 1.00 (Ref.) | 63 974 | 1.7 | 1.00 (Ref.) | 22 626 | 0.6 | 1.00 (Ref.) |
| Yes | 1044 | 5.4 | 1.29 (1.21-1.37) | 462 | 2.3 | 1.21 (1.10-1.23) | 132 | 0.7 | 1.06 (0.90-1.26) |
| Subtypes of maternal CVD<sup>c</sup> |  |  |  |  |  |  |  |  |  |
| Cerebrovascular disease | 204 | 6.0 | 1.40 (1.22-1.61) | 99 | 2.9 | 1.54 (1.26-1.88) | 28 | 0.8 | 1.30 (0.90-1.88) |
| Arrhythmia | 565 | 5.2 | 1.24 (1.14-1.34) | 242 | 2.2 | 1.11 (0.97-1.26) | 57 | 0.5 | 0.81 (0.63-1.05) |
| Heart failure | 26 | 6.4 | 2.04 (1.08-2.34) | 18 | 4.4 | 2.12 (1.30-3.46) | <5<sup>l</sup> | 0.9 | 1.54 (0.58-4.11) |
| Valvular heart disease | 85 | 4.9 | 1.15 (0.93-1.43) | 41 | 2.3 | 1.20 (0.88-1.65) | 19 | 1.1 | 1.73 (1.10-2.71) |
| Congenital heart disease | 239 | 5.4 | 1.35 (1.19-1.54) | 108 | 2.4 | 1.24 (1.02-1.51) | 35 | 0.8 | 1.26 (0.90-1.75) |
| **Paternal CVD (N=2 672 229)** |  |  |  |  |  |  |  |  |  |
| Composite paternal CVD |  |  |  |  |  |  |  |  |  |
| No | 151 794 | 4.1 | 1.00 (Ref.) | 63 281 | 1.7 | 1.00 (Ref.) | 22 367 | 0.6 | 1.00 (Ref.) |
| Yes | 1212 | 4.7 | 1.10 (1.04-1.16) | 572 | 2.2 | 1.13 (1.04-1.22) | 185 | 0.7 | 1.13 (0.98-1.30) |
| Subtypes of paternal CVD<sup>c</sup> |  |  |  |  |  |  |  |  |  |
| Cerebrovascular disease | 254 | 5.8 | 1.36 (1.20-1.54) | 103 | 2.3 | 1.22 (1.00-1.48) | 34 | 0.8 | 1.24 (0.88-1.74) |
| Arrhythmia | 710 | 4.3 | 1.02 (0.94-1.09) | 349 | 2.1 | 1.09 (0.98-1.21) | 100 | 0.6 | 0.96 (0.79-1.17) |
| Heart failure | 55 | 5.2 | 1.23 (0.94-1.60) | 24 | 2.3 | 1.14 (0.77-1.70) | 11 | 1.0 | 1.62 (0.90-2.93) |
| Valvular heart disease | 121 | 4.2 | 0.98 (0.82-1.17) | 58 | 2.0 | 1.03 (0.80-1.34) | 24 | 0.8 | 1.33 (0.89-1.98) |
| Congenital heart disease | 185 | 4.9 | 1.20 (1.04-1.39) | 96 | 2.5 | 1.30 (1.06-1.58) | 29 | 0.7 | 1.21 (0.84-1.74) |
| British Columbia, Canada |  |  |  |  |  |  |  |  |  |
| **Maternal CVD (N=887 582)** |  |  |  |  |  |  |  |  |  |
| Composite maternal CVD |  |  |  |  |  |  |  |  |  |
| No | 76 290 | 7.0 | 1.00 (Ref.) | 18 404 | 1.6 | 1.00 (Ref.) | 4035 | 0.3 | 1.00 (Ref.) |
| Yes | 2043 | 9.5 | 1.29 (1.23-1.35) | 614 | 2.7 | 1.28 (1.17-1.39) | 97 | 0.4 | 1.18 (0.96-1.44) |
| Subtypes of maternal CVD<sup>c</sup> |  |  |  |  |  |  |  |  |  |
| Cerebrovascular disease | 195 | 10.1 | 1.41 (1.21-1.64) | 57 | 2.8 | 1.29 (0.95-1.74) | 13 | 0.6 | 1.79 (1.04-3.09) |
| Arrhythmia | 1274 | 9.4 | 1.25 (1.18-1.33) | 407 | 2.9 | 1.29 (1.16-1.43) | 63 | 0.4 | 1.21 (0.95-1.56) |
| Heart failure | 68 | 10.9 | 1.47 (1.13-1.91) | 19 | 2.9 | 1.22 (0.75-1.98) | <5<sup>d</sup> | 0.6 | 1.65 (0.62-4.40) |
| Valvular heart disease | 300 | 8.2 | 1.16 (1.03-1.31) | 73 | 1.9 | 1.22 (0.95-1.56) | 9 | 0.2 | 0.65 (0.34-1.25) |
| Congenital heart disease | 390 | 10.5 | 1.44 (1.29-1.61) | 117 | 3.0 | 1.30 (1.07-1.59) | 18 | 0.5 | 1.28 (0.80-2.02) |

| **Paternal CVD (N=811 481)** |  |  |  |  |  |  |  |  |  |
| --- | --- | --- | --- | --- | --- | --- | --- | --- | --- |
| Composite paternal CVD |  |  |  |  |  |  |  |  |  |
| No | 71 270 | 7.0 | 1.00 (Ref.) | 16 911 | 1.6 | 1.00 (Ref.) | 3778 | 0.4 | 1.00 (Ref.) |
| Yes | 1066 | 8.0 | 1.07 (1.01-1.14) | 322 | 2.3 | 1.06 (0.94-1.19) | 52 | 0.4 | 1.03 (0.78-1.36) |
| Subtypes of paternal CVD<sup>c</sup> |  |  |  |  |  |  |  |  |  |
| Cerebrovascular disease | 150 | 9.5 | 1.29 (1.09-1.52) | 57 | 3.4 | 1.58 (1.21-2.06) | 9 | 0.5 | 1.51 (0.78-2.90) |
| Arrhythmia | 677 | 7.4 | 0.99 (0.91-1.07) | 219 | 2.3 | 1.05 (0.92-1.21) | 27 | 0.3 | 0.78 (0.53-1.14) |
| Heart failure | 61 | 9.3 | 1.31 (1.00-1.73) | 16 | 2.3 | 1.11 (0.68-1.81) | <5 | 0.3 | 0.83 (0.21-3.30) |
| Valvular heart disease | 125 | 8.4 | 1.10 (0.91-1.33) | 33 | 2.1 | 1.05 (0.73-1.50) | 7 | 0.4 | 1.21 (0.58-2.53) |
| Congenital heart disease | 196 | 9.0 | 1.23 (1.06-1.42) | 47 | 2.0 | 0.84 (0.62-1.14) | 10 | 0.4 | 1.24 (0.63-2.43) |

Note: ADHD, attention-deficit/ hyperactivity disorder; ASD, autism spectrum disorder; CI, confidence interval; CVD, cardiovascular disease; HR, hazard ratio; ID, intellectual disability.
<sup>a</sup>Incidence rates per 1000 child-years.
<sup>b</sup>The HRs were minimally adjusted for child's age, birthyear, and sex.
<sup>c</sup>All CVD subtypes were coded as binary: No (reference)/Yes.
<sup>d</sup>Exact count is suppressed for confidentiality reason.

Figure 1 shows the HRs fully adjusted for maternal and offspring characteristics for each country and for the pooled analysis. The meta-analysis suggested that maternal CVD, overall, was associated with 15% higher hazard of ADHD (HR: 1.15; 95% CI: 1.10-1.20; I-squared: 0%; P for heterogeneity: .54) and 15% higher hazard of ASD (HR: 1.15; 95% CI: 1.02-1.23; I-squared: 0%; P for heterogeneity: .50) in offspring, when compared with no maternal CVD. However, no association was found between maternal CVD and ID in offspring (HR: 1.03; 95% CI: 0.89-1.16; I-squared: 0%; P for heterogeneity: 0.53).

**Fig. 1.**
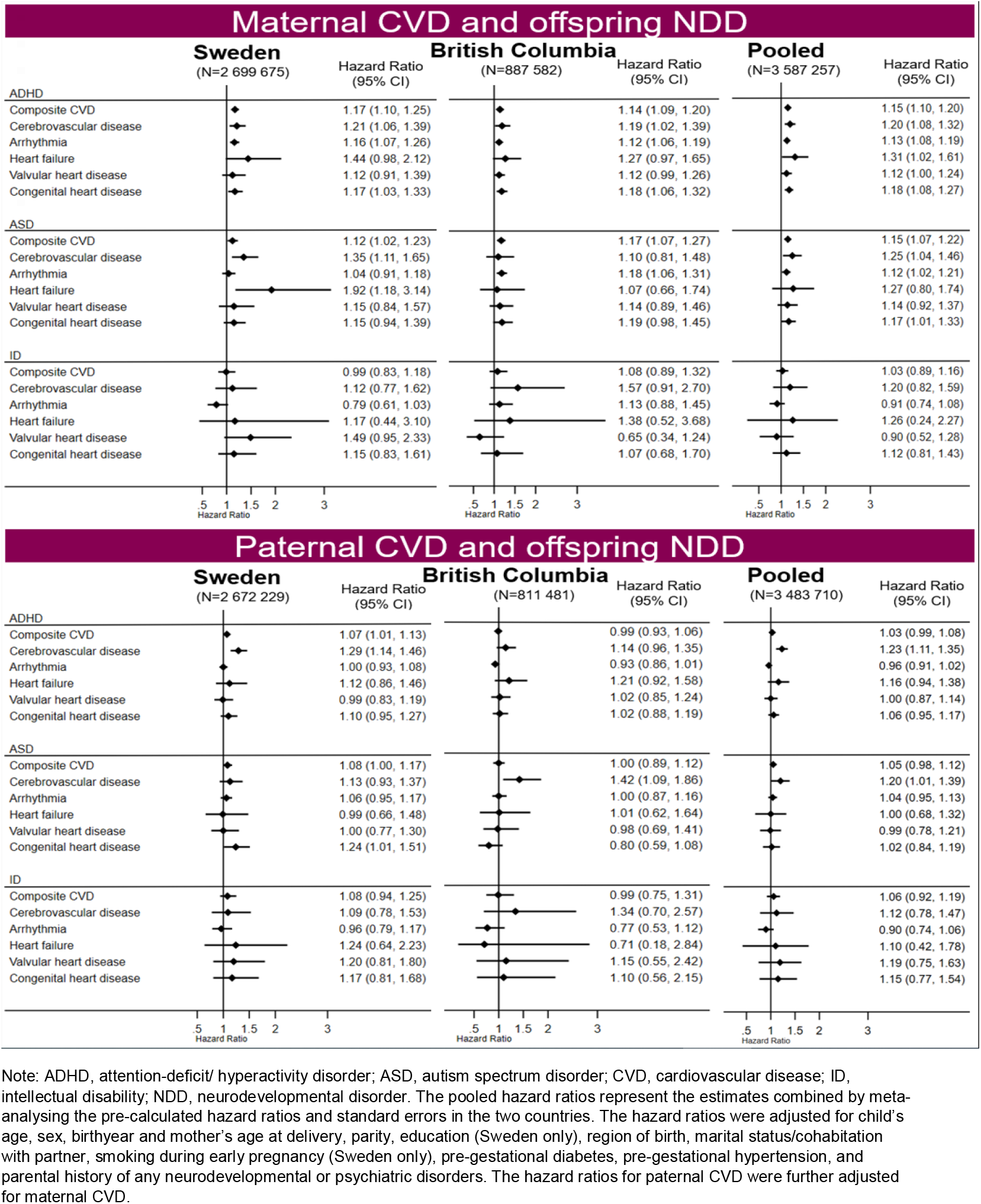
Plots showing adjusted hazard ratios of neurodevelopmental disorders according to pre-existing maternal and paternal cardiovascular diseases: singleton offspring live-born without major malformations in Sweden 1990 to 2019 and in British Columbia, Canada 1992 to 2019.

Among the subtypes of maternal CVD, increased hazard of ADHD in offspring was found for all included subtypes, namely maternal heart failure (HR 1.31; 95% CI: 1.02-1.61), cerebrovascular disease (HR 1.20; 95% CI: 1.08-1.32), congenital heart disease (HR 1.18; 95% CI: 1.08-1.27), arrhythmia (HR 1.13; 95% CI: 1.08-1.19), and valvular heart disease (HR 1.12; 95% CI: 1.00-1.24) when compared with offspring of mothers without the respective CVD condition. Offspring exposed to maternal cerebrovascular disease, congenital heart disease, and arrythmia had 1.25 (95% CI: 1.04-1.46), 1.17 (95% CI: 1.01-1.33), and 1.12 (95% CI: 1.01-1.21) times elevated HRs of ASD, respectively, compared with non-exposed offspring. None of the maternal CVD subtypes were associated with ID in offspring (Figure 1).

The paternal comparison models suggested no associations between paternal CVD and rates of ADHD, ASD or ID in offspring (Figure 1). However, when the analysis was stratified by subtypes of paternal CVD, offspring exposed to paternal cerebrovascular disease showed increased hazard of ADHD (HR 1.23; 95% CI 1.11-1.35) and ASD (HR 1.20; 95% CI 1.01-1.39).

The natural indirect effects in the causal mediation analysis (eTable2) indicated that the total effects of maternal CVD on ADHD and ASD in offspring were not considerably mediated through preterm birth, and about 90% of the total effect was explained through other undiscovered pathways.

In the sensitivity analyses, adjustment for maternal BMI in Sweden (eTable 4) or parental neighborhood income quintiles in BC (eTable 5) did not influence the results. Furthermore, multiple imputation of missing data (eTable 6) and analyses restricted to later-born cohorts (eTable 7, eTable 8) yielded results consistent with those obtained in the main analyses. The estimated associations between maternal CVDs and offspring ASD were robust to the exclusion of ID diagnoses (eTable 9). The pattern of associations in the paternal comparison models remained largely unchanged when paternal age at childbirth was adjusted for (eFigure 3).

## Discussion

In this large-scale cohort study including nearly 3.6 million children born in Sweden or BC, Canada, we found that pre-pregnancy maternal CVD was associated with increased risks of ADHD and ASD in offspring. The causal mediation analyses suggested that the observed associations were largely independent of preterm delivery. We found no excess risk of ID among offspring exposed to maternal CVD. Paternal CVD was not generally associated with offspring risks of NDD outcomes.

To our knowledge, this is the first population-based study to report the risk of NDDs in children of mothers with pre-existing cardiovascular conditions. Our results are broadly consistent with the existing literature documenting elevated risks of NDDs in children exposed to maternal cardiovascular risk factors including obesity^14^, hypertension^19,20^, and diabetes^18^. A meta-analysis reported higher risks of ASD and ADHD among children exposed to maternal hypertensive disorders, although most of the included studies did not adequately control for confounding^20^.

The association of maternal CVD with offspring risk of ADHD and ASD in the current study persisted after adjustment for parental history of any neurodevelopmental or psychiatric disorders and a large number of other covariates related to maternal social and health characteristics. This association is unlikely to be explained by shared genetic liability given the general absence of such association with paternal CVD, although an exception may be the association with cerebrovascular disease, that could be attributable to genetic influences. Our results thus broadly suggest that the maternal intra-uterine environment plays a greater role than genetics in the pathophysiological mechanism behind the association between CVD and NDDs. We found very little mediating effect of preterm birth, a finding in line with an investigation showing that the increased risk of ASD and ADHD in children of mothers with hypertensive disorders during pregnancy was independent from fetal growth and gestational age at birth^19^.

The precise mechanisms through which a pre-existing cardiovascular dysfunction in mothers can influence subsequent disease risks in children are not clearly understood. Pre-conceptional CVD in women may be accompanied by a range of physiological disturbances during pregnancy, including abnormal uteroplacental perfusion, perturbation of hormonal homeostasis, metabolic dysfunction, increased inflammation, and oxidative stress^5,6^ that may adversely affect the structural brain development in offspring. Impaired placentation can be hypothesized to be a major candidate pathway since placental perfusion is primarily a function of cardiac performance during pregnancy. Available evidence suggests a link between pre-existing cardiac dysfunction and poor placental circulation which in turn contributes to pregnancy-related and neonatal complications^37,38^. CVD medication use during pregnancy is another possible mediating mechanism, since brain development of the fetus is particularly vulnerable to maternal medication exposure^39^. Future research aiming to explore the mechanisms could benefit from studying more specific maternal CVDs rather than broad diagnostic categories.

### Strengths and Limitations

The strengths of our study include the inclusion of data from several linked population-based registers in two countries, with almost 30 years of follow-up. This allowed us to estimate and cross-validate the associations between a broad spectrum of maternal cardiovascular conditions and long-term neurodevelopmental outcomes in offspring. In addition to adjusting for several important maternal confounders, including smoking and neurodevelopmental and psychiatric conditions, we adopted a family-based study design to explore the presence of unmeasured genetic confounding. The Swedish population registers are generally known to have high quality and nation-wide coverage^24,25^, minimizing selection bias due to systematic losses to follow up. On the other hand, the BC cohort is known to be ethnically diverse^40^ and is more heterogeneous than the Swedish cohort, enhancing the generalizability of the findings.

Some limitations of the study should be acknowledged. First, although we adjusted for parental history of any neurodevelopmental or psychiatric comorbidities in the analysis, residual confounding may still remain due to possible underdiagnoses of parents’ NDD. Second, while the study had adequate statistical power to estimate the associations between overall maternal CVD and the outcomes, there was insufficient number of cases in some strata of cardiovascular conditions, which might obscure true associations, particularly with ID. Third, as data on maternal cardiovascular conditions and neurodevelopmental outcomes in Sweden came from specialist settings, and outpatient diagnoses were only available from 2001 onward, milder cases were less likely to be captured. However, our BC data came from both specialized and primary care settings and we further used data from drug registers for identification of ADHD cases. Moreover, increased awareness and improvements in diagnostic criteria over time imply that the exposure and outcomes are more prone to be misclassified in the early years of follow up. However, such potential misclassifications are expected to be nondifferential and would result in underestimation of the true effect sizes.

## Conclusion

This large population-based cohort study revealed that prexisting maternal CVD might be a risk factor for ADHD and ASD in offspring. The observed risks appear to be unexplained by shared genetic factors, suggesting the involvement of intrauterine mechanisms. While the precise mechanisms need to be carefully assessed in future studies, the findings highlight the importance of close clinical monitoring and support to reproductive aged women with an undelying CVD for prevention of ADHD and ASD in children.

## Data Availability

All data produced in the present work are contained in the manuscript

## Acknowledgements

The authors would like to thank Jonas Söderling at Karolinska Institute for his assistance with data management and Emma Wenn at the University of British Columbia for her assistance with data management and analysis.

## Author Contributions

Drs Razaz and Hossin had access to the data used in the study and take responsibility for the integrity of the data and the accuracy of the data analysis.

*Concept and design:* Hossin, Razaz

*Acquisition, analysis, or interpretation of data:* All authors

*Drafting of the manuscript:* Hossin

*Critical revision of the manuscript for important intellectual content:* All authors

*Statistical analysis:* Hossin

*Obtained funding:* Razaz

*Administrative, technical, or material support:* Razaz

*Supervision:* Razaz

