## Supplementary for "Association of pre-existing maternal cardiovascular diseases with neurodevelopmental disorders in offspring: a cohort study in Sweden and British Columbia, Canada"

**eFigure 1.** Flow diagram of the study samples

**eFigure 2.** A conceptual framework of the study

**eFigure 3.** Hazard ratios of neurodevelopmental disorders according to paternal cardiovascular disease, adjusted for paternal age at child's birth: singleton offspring live-born without major malformations in Sweden 1990 to 2019 (N=2 672 229) and British Columbia, Canada 1992 to 2019 (N=741 026)

**eTable 1.** International Classification of Disease (ICD) codes used to define maternal diseases and offspring neurodevelopmental disorders

**eTable 2.** Register databases from which the study variables were derived

**eTable 3.** Causal mediation analysis to estimate the impact of preterm delivery on the association between maternal pre-existing cardiovascular disease and offspring's neurodevelopmental disorders: singleton offspring live-born without major malformations in Sweden 1990 to 2019 (N=2 672 229) and British Columbia, Canada 1992 to 2019 (N= 887 582).

**eTable 4.** Incidence rates and hazard ratios of neurodevelopmental disorders according to pre-existing maternal cardiovascular diseases, adjusted for maternal early pregnancy body mass index: singleton offspring live-born without major malformations in Sweden 1992 to 2019 (N=2 311 311).

**eTable 5.** Incidence rates and hazard ratios of neurodevelopmental disorders according to pre-existing maternal cardiovascular diseases, adjusted for parental neighbourhood income quintiles: singleton offspring live-born without major malformations in British Columbia, Canada 1992 to 2019 (N= 863 440)

**eTable 6.** Multiple-imputation analysis of the associations between maternal pre-existing cardiovascular diseases and offspring's neurodevelopmental disorders: singleton offspring live-born without major malformations in Sweden 1990 to 2019 (N=2 940 626).

**eTable 7.** Sensitivity analysis restricted to later-born cohorts, concerning the associations between maternal pre-existing cardiovascular diseases and offspring's neurodevelopmental disorders: singleton offspring live-born without major malformations in Sweden 1997 to 2019 (N=1 960 441).

**eTable 8.** Sensitivity analysis restricted to later-born cohorts, concerning the associations between pre-existing maternal cardiovascular diseases and offspring's neurodevelopmental disorders: singleton offspring live-born without major malformations in British Columbia, Canada 2001 to 2019 (N= 589 933).

**eTable 9.** Incidence rates and hazard ratios of autism spectrum disorder without intellectual disability according to pre-existing maternal cardiovascular diseases:

singleton offspring live-born without major malformations in Sweden 1990 to 2019 (N=2 672 229) and British Columbia, Canada 1992 to 2019 (N= 887 582).

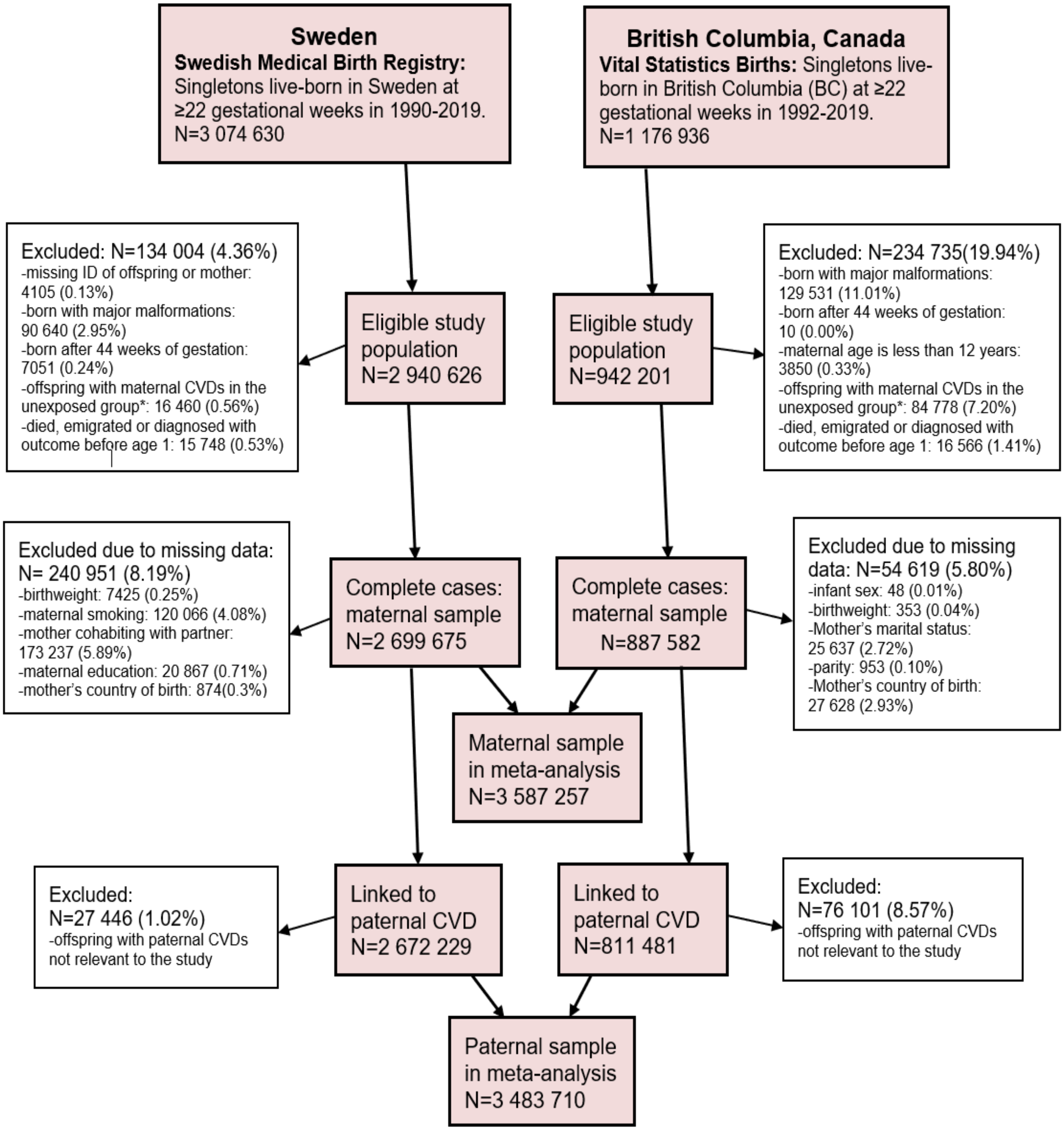


*Excluded from the unexposed group were the offspring of mothers diagnosed with any cardiovascular diseases, except those with the diagnoses for hypertensive diseases.

**eFigure 1. Flow diagram of the study samples.**

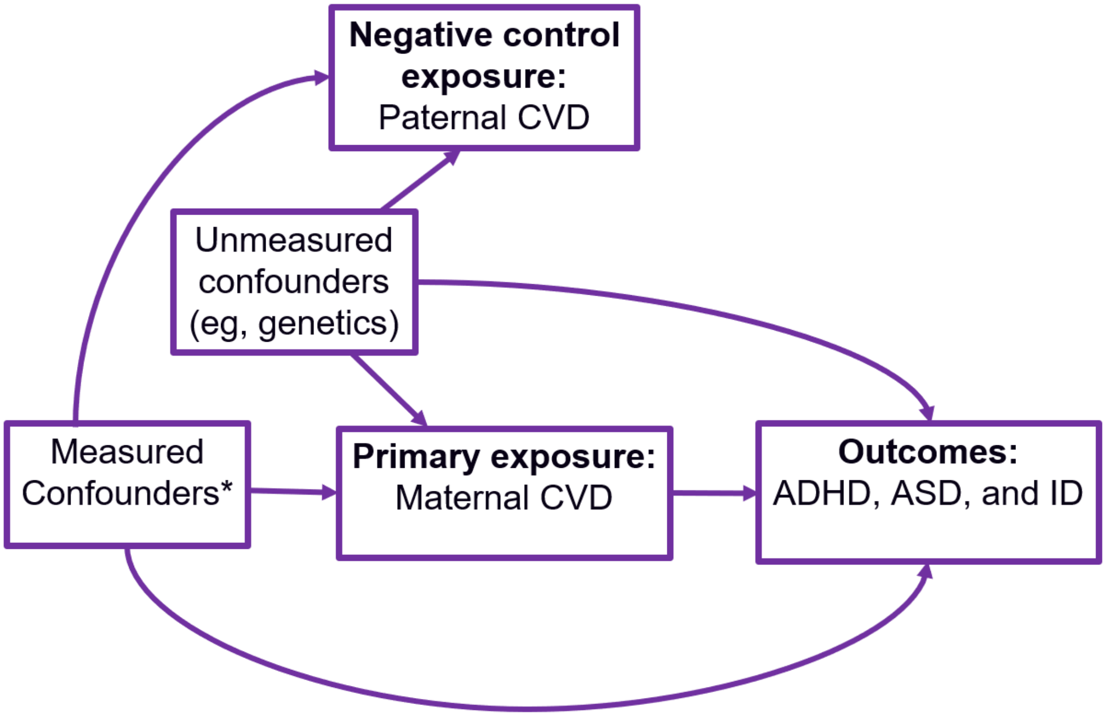


Note: ADHD, attention-deficit/hyperactivity disorder; ASD, autism spectrum disorder; CVD, cardiovascular disease; ID, intellectual disability.
*The measured confounders include mother's age at delivery, parity, education, country of birth, marital/cohabitation status, smoking during early pregnancy, pre-gestational diabetes, pre-gestational hypertension, and parental history of any neurodevelopmental or psychiatric disorders.

**eFigure 2. A conceptual framework of the study**

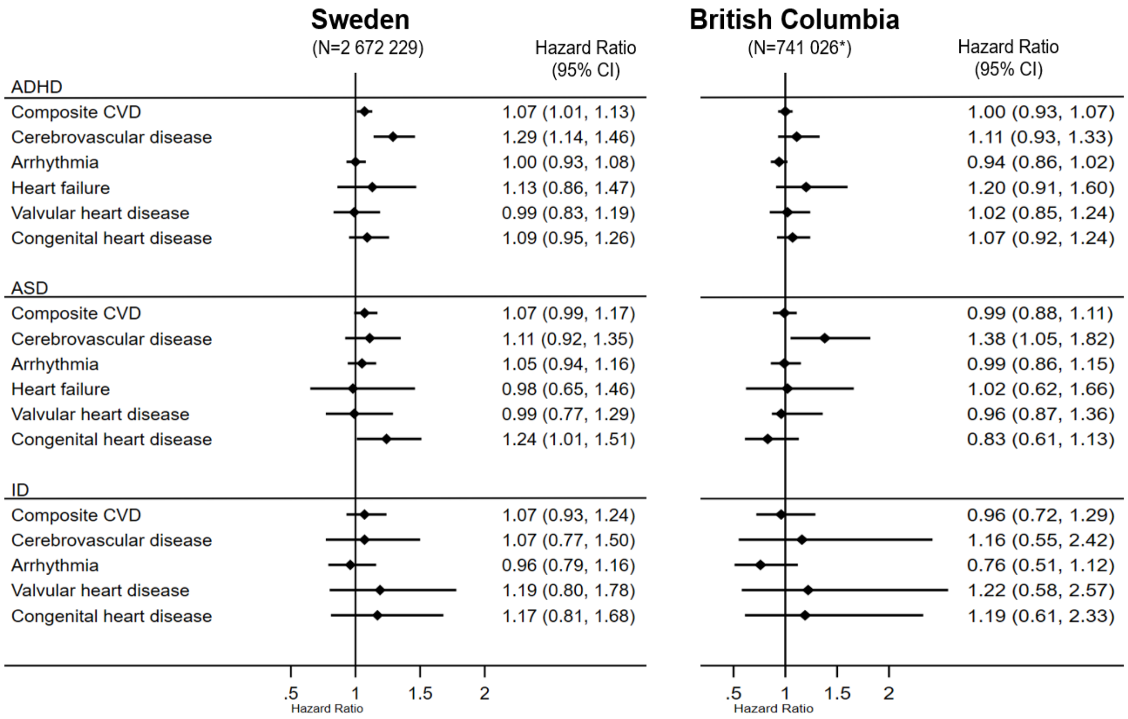


*Offspring with missing data on paternal age were excluded from BC data. The Swedish data had no missing on paternal age.

Note: ADHD, attention-deficit/hyperactivity disorder; ASD, autism spectrum disorder; CVD, cardiovascular disease; ID, intellectual disability. All hazard ratios were adjusted for child's age, sex, birthyear; paternal age at delivery; and mother's CVD, age at delivery, parity, education, country of birth, cohabitation with partner, smoking during pregnancy, pre-gestational diabetes, pre-gestational hypertension, and parental history of any neurodevelopmental or psychiatric disorders.

**eFigure 3. Hazard ratios of neurodevelopmental disorders according to paternal cardiovascular disease, adjusted for paternal age at child's birth: singleton offspring live-born without major malformations in Sweden 1990 to 2019 (N=2 672 229) and British Columbia, Canada 1992 to 2019 (N= 741 026)**

**eTable 1. International Classification of Disease (ICD) codes used to define maternal diseases and offspring neurodevelopmental disorders**

| | ICD-9 | ICD-10 |
|---|---|---|
| **Maternal diseases** | | |
| Cerebrovascular disease | 430-438 | I60-I69, G45 |
| Arrhythmia | 426-427 | I44-I49 |
| Heart failure | 428 | I50 |
| Valvular heart disease (non-congenital) | 394-397, 424 | I05-I09, I34-I37 |
| Congenital heart disease | 745-747 | Q20-Q26 |
| Any psychiatric disorders | 290-319 | F00-F99 |
| Neurodevelopmental disorders | 299, 314, 317-319 | F84, F70-F73, F78-F79, F90 |
| Pre-gestational diabetes | 250, 648A | E10-E14, O24.1-O24.3 |
| Pre-gestational hypertension* | 401-405, 642C, 642H | I10-I15, O10-O11 |
| **Offspring neurodevelopmental disorders** | | |
| ADHD[#] | 314 | F90 |
| ASD | 299 | F84 |
| ID | 317-319 | F70-F73, F78-F79 |

ADHD, attention-deficit/ dyperactivity disorder; ASD, autism spectrum disorder; ICD, international classification of disease; ID, intellectual disability.

*Pre-gestational hypertension was also self-reported in a checkbox at the first prenatal visit.

[#]The ADHD cases were additionally identified through dispensed drugs using the following Anatomical Therapeutic Chemical (ATC) codes, available since 2005 in Sweden and since 1996 in British Columbia: N06BA01 (amphetamine), N06BA02 (dexamfetamine), N06BA04 (methylphenidate), N06BA09 (atomoxetine), N06BA12 (lisdexamfetamine).

**eTable 2. Register databases from which the study variables were derived**

| Major variables in the study | Register data sources* | |
|---|---|---|
| | Sweden | BC, Canada |
| **Neonatal characteristics** | | |
| Birth year | Medical Birth Register | Vital Statistics Births |
| Sex | Medical Birth Register | Vital Statistics Births |
| Preterm birth (<37 gestational weeks) | Medical Birth Register | Vital Statistics Births |
| Small for gestational age (<10th percentile) | Medical Birth Register | Vital Statistics Births |
| ADHD | National Patient Register<br>Prescribed Drug Register | Discharge Abstract Database<br>Medical Services Plan<br>PharmaNet |
| ASD | National Patient Register | Discharge Abstract Database<br>Medical Services Plan |
| ID | National Patient Register | Discharge Abstract Database<br>Medical Services Plan |
| **Maternal characteristics** | | |
| Age at delivery | Medical Birth Register | Vital Statistics Births |
| Parity | Medical Birth Register | Vital Statistics Births |
| Marital/cohabitation status | Medical Birth Register | Vital Statistics Births |
| Region of birth | Total Population Register | Vital Statistics Births |
| Education | Education Register | Not available |
| Smoking during early pregnancy | Medical Birth Register | Not available |
| Pre-existing CVD | National Patient Register | Discharge Abstract Database<br>Medical Services Plan |
| Neurodevelopmental and psychiatric disorders | National Patient Register | Discharge Abstract Database<br>Medical Services Plan |
| Pre-gestational hypertension | National Patient Register<br>Medical Birth Register | Discharge Abstract Database<br>Medical Services Plan |
| Pre-pregnancy diabetes | National Patient Register<br>Medical Birth Register | Discharge Abstract Database<br>Medical Services Plan |

Note: ADHD, attention-deficit/hyperactivity disorder; ASD, autism spectrum disorder; CVD, cardiovascular disease; ID, intellectual disability.

*A detailed description of all register databases used in the study is given below:

<u>The Swedish Medical Birth Register</u>

The Swedish Medical Birth Register (MBR) retrieves data on prenatal, obstetric, and neonatal characteristics, including maternal diagnoses, based on medical records from the prenatal care, the delivery care, and the neonatal care. Founded in 1973, the MBR covers >98% of all births occurring in Sweden[1].

<u>The Swedish National Patient Register</u>

The National Patient Register (NPR) consists of the inpatient and outpatient registers and holds data on health care from both public and private caregivers. The inpatient register, also known as hospital discharge register, was initiated in 1964 (psychiatric diagnoses since 1973), but complete nationwide coverage was obtained in 1987. The outpatient register was launched in 2001 and records information on visits to specialized outpatient care. The diagnoses of diseases recorded in the NPR are identified using the Swedish version of the International Classification of Disease (ICD) codes[2].

<u>The Swedish Cause of Death Register</u>

The Swedish Cause of Death Register (CDR) is an excellent source of data for register-based research and contains information on all deaths in Sweden since 1952. The register is known for its high quality and complete data and is linked to other national registers through unique personal identification numbers[3]. Unlike the NPR that uses the Swedish version of ICD codes to classify diseases, the CDR uses the international version of the classification system to facilitate international comparison of cause-specific mortality statistics[4].

The Swedish Total Population Register

Since 1968, the Total Population Register (TPR) contains sociodemographic information (eg, sex, country of birth) as well as data on important life events including dates of birth, death and migration of all residents in Sweden. The TPR allows complete follow up and censoring of individuals and thereby minimizes the risk of selection bias[5].

The Swedish Education Register

Since 1985, the Education Register in Sweden contains annually updated data on completed education, collected from more than 30 different sources including the schools and education providers and questionnaire surveys. Data on education is coded according to the Swedish education nomenclature SUN, a classification system which was later adapted to the International Standard Classification of Education (ISCED 97) in 2000, distinguishing between the level and type of education[6]

The Swedish Prescribed Drug Register

The Prescribed Drug Register (PDR) was established in July 2005 and contains data on all outpatient prescribed drugs dispensed at pharmacies throughout Sweden. The data on the drug prescriptions recorded in the PDR is retrieved from the Swedish eHealth Agency. The overall quality of the data in the register is good, with minimal errors and incompletion. The World Health Organization's Anatomical Therapeutic Chemical (ATC) classification system is used to classify all drugs in the PDR[7].

The BC Vital Statistics

The BC Vital Statistics database contains both birth and death files. The BC Vital Statistics Birth database covers >98% of all births in British Columbia (BC) and contains information on maternal characteristics, diseases, pregnancy, delivery and neonatal complications, and birth characteristics from January 1, 1985[8]. On the other hand, information on all deaths occurring in BC since January 1, 1985 is retrieved from the BC Vital Statistics Death file[9].

The BC Discharge Abstract Database

The Discharge Abstract Database (DAD) captures administrative, demographic and clinical data of in-patients in BC since April 1, 1985, including information of hospital admission and discharge dates, day surgery, and diagnoses using ICD-9/10 codes[10].

The BC Medical Services Plan

Established in 1965, the BC Medical Services Plan (MSP) a provincial health insurance program covering all eligible residents in BC. The MSP files contain both specialist and primary care physician billings data covering all fee-for-service outpatient claims, including service date and associated diagnoses based on ICD-9 codes from April 1, 1991 onwards[11].

The BC PharmaNet

PharmaNet is province-wide electronic medical record system containing information on all outpatient prescriptions dispensed from community pharmacies in BC from January 1,

1996[12]. Information includes the dispensation date, drug name or unique Drug Identification Numbers, which are mapped to ATC classification system.

The BC Central Demographics File

The Central Demographics File, previously known as the Consolidation File, provides basic demographic (eg, age and sex) and registration data for provincial health coverage from January 1, 1986 onward.

**eTable 3. Causal mediation analysis to estimate the impact of preterm delivery on the association between maternal pre-existing cardiovascular disease and offspring's neurodevelopmental disorders: singleton offspring live-born without major malformations in Sweden 1990 to 2019 (N=2 672 229) and British Columbia, Canada 1992 to 2019 (N= 887 582).**

| Parameters[#] | ADHD | ASD |
|---|---|---|
| | IRR (95% CI) | HR (95% CI) |
| **Sweden** | | |
| Total Effect (TE) | 1.16 (1.09-1.23) | 1.11 (1.01-1.21) |
| Natural Direct Effect (NDE)[1] | 1.15 (1.08-1.23) | 1.09 (1.00-1.20) |
| Natural Indirect Effect (NIE) | 1.01 (1.00-1.01) | 1.01 (1.00-1.02) |
| Controlled Direct Effect (CDE)[2] | 1.35 (1.11-1.65) | 1.51 (1.16-1.96) |
| Mediated proportion[3] | 5% | 10% |
| **British Columbia, Canada** | | |
| Total Effect (TE) | 1.13 (1.08-1.18) | 1.17 (1.07-1.26) |
| Natural Direct Effect (NDE) [1] | 1.13 (1.08-1.18) | 1.17 (1.07-1.26) |
| Natural Indirect Effect (NIE) | 1.00 (1.00-1.00) | 1.00 (1.00-1.00) |
| Controlled Direct Effect (CDE)[2] | 1.13 (1.08-1.18) | 1.18 (1.08-1.28) |
| Mediated proportion[3] | 0% | 0% |

Note: ADHD, attention-deficit/hyperactivity disorder; ASD, autism spectrum disorder; CI, confidence interval; CVD, cardiovascular disease; IRR, incidence rate ratios.

#All parameters were derived from Poisson regression models and were conditional on the following covariates: child's age, birth year, and sex; mother's age at delivery, parity, education (Sweden only), region of birth, marital status/cohabitation with partner, smoking during early pregnancy (Sweden only), pre-gestational diabetes, pre-gestational hypertension, and parental history of any neurodevelopmental or psychiatric disorders.

[1]The NDE represents the effect of maternal CVD in the absence of preterm birth.

[2]The CDE represents the effect of maternal CVD obtained by setting the value of preterm birth to 1 (i.e., everyone is assumed to be preterm).

[3]The proportion mediated was calculated using the formula: $\{IRR^{NDE} (IRR^{NIE} - 1)/(IRR^{NDE} * IRR^{NIE} - 1)\}*100$.

**eTable 4. Incidence rates and hazard ratios of neurodevelopmental disorders according to pre-existing maternal cardiovascular diseases, adjusted for maternal early pregnancy body mass index: singleton offspring live-born without major malformations in Sweden 1992 to 2019 (N=2 311 311#).**

| Exposure/s | Outcomes | | | | |
|---|---|---|---|---|---|
| | **ADHD** | | | | |
| | No. of events | Rates* | Model 1 HR (95% CI) | Model 2 HR (95% CI) | Model 3 HR (95% CI) |
| Composite maternal CVD | | | | | |
| No | 126 305 | 4.4 | 1.00 (Reference) | 1.00 (Reference) | 1.00 (Reference) |
| Yes | 945 | 5.4 | 1.28 (1.20-1.36) | 1.16 (1.09-1.23) | 1.17 (1.10-1.24) |
| Subtypes of maternal CVD | | | | | |
| Cerebrovascular disease | 178 | 5.9 | 1.37 (1.18-1.58) | 1.18 (1.01-1.36) | 1.17 (1.01-1.36) |
| Arrhythmia | 519 | 5.2 | 1.24 (1.14-1.35) | 1.16 (1.06-1.27) | 1.17 (1.07-1.28) |
| Heart failure | 24 | 6.6 | 1.62 (1.09-2.42) | 1.44 (0.97-2.15) | 1.39 (0.93-2.07) |
| Valvular heart disease | 76 | 4.8 | 1.12 (0.90-1.41) | 1.09 (0.87-1.37) | 1.10 (0.88-1.38) |
| Congenital heart disease | 212 | 5.3 | 1.25 (1.08-1.45) | 1.12 (0.98-1.28) | 1.13 (0.99-1.30) |
| | **ASD** | | | | |
| Composite maternal CVD | | | | | |
| No | 53 337 | 1.8 | 1.00 (Reference) | 1.00 (Reference) | 1.00 (Reference) |
| Yes | 424 | 2.4 | 1.20 (1.09-1.32) | 1.12 (1.01-1.23) | 1.12 (1.02-1.24) |
| Subtypes of maternal CVD | | | | | |
| Cerebrovascular disease | 84 | 2.8 | 1.41 (1.14-1.75) | 1.25 (1.01-1.55) | 1.25 (1.0-1.55) |
| Arrhythmia | 228 | 2.3 | 1.13 (0.99-1.29) | 1.07 (0.94-1.22) | 1.08 (0.95-1.23) |
| Heart failure | 17 | 4.6 | 2.32 (1.45-3.74) | 2.08 (1.29-3.34) | 2.00 (1.24-3.22) |
| Valvular heart disease | 37 | 2.3 | 1.17 (0.85-1.61) | 1.11 (0.81-1.53) | 1.12 (0.81-1.54) |
| Congenital heart disease | 97 | 2.4 | 1.21 (0.99-1.48) | 1.012(0.92-1.37) | 1.13 (0.93-1.38) |
| | **ID** | | | | |
| Composite maternal CVD | | | | | |
| No | 17 927 | 0.6 | 1.00 (Reference) | 1.00 (Reference) | 1.00 (Reference) |
| Yes | 121 | 0.7 | 1.07 (0.90-1.28) | 1.00 (0.84-1.20) | 1.01 (0.84-1.21) |
| Subtypes of maternal CVD | | | | | |
| Cerebrovascular disease | 25 | 0.8 | 1.31 (0.88-1.94) | 1.14 (0.77-1.69) | 1.15 (0.77-1.70) |
| Arrhythmia | 52 | 0.5 | 0.81 (0.62-1.06) | 0.79 (0.60-1.04) | 0.80 (0.61-1.05) |
| Heart failure | <5 | 1.1 | - | - | - |
| Valvular heart disease | 16 | 1.0 | 1.59 (0.97-2.59) | 1.37 (0.84-2.24) | 1.37 (0.84-2.25) |
| Congenital heart disease | 32 | 0.8 | 1.25 (0.88-1.77) | 1.15 (0.81-1.63) | 1.16 (0.83-1.65) |

Note: ADHD, attention-deficit/hyperactivity disorder; ASD, autism spectrum disorder; CI, confidence interval; CVD, cardiovascular disease; HR, hazard ratio; ID, intellectual disability.
#Children with missing data on maternal body mass index were excluded.
*Incidence rates per 1000 child-years.
Model 1 was minimally adjusted for child's age, birth year and sex. Model 2 was further adjusted for mother's age at delivery, parity, education, region of birth, cohabitation with partner, smoking during early pregnancy, pre-gestational diabetes, pre-gestational hypertension, and parental history of any neurodevelopmental or psychiatric disorders. Model 3 was additionally adjusted for maternal early pregnancy body mass index.
-Not estimated due to insufficient events

**eTable 5. Incidence rates and hazard ratios of neurodevelopmental disorders according to pre-existing maternal cardiovascular diseases, adjusted for parental neighbourhood income quintiles: singleton offspring live-born without major malformations in British Columbia, Canada 1992 to 2019 (N= 863 440).**

| Exposure/s | Outcomes | | | | |
|---|---|---|---|---|---|
| | **ADHD** | | | | |
| | No. of events | Rates* | Model 1 HR (95% CI) | Model 2 HR (95% CI) | Model 3 HR (95% CI) |
| Composite maternal CVD | | | | | |
| No | 75 859 | 7.2 | 1.00 (Reference) | 1.00 (Reference) | 1.00 (Reference) |
| Yes | 2037 | 9.6 | 1.28 (1.22-1.34) | 1.14 (1.09-1.20) | 1.14 (1.09-1.20) |
| Subtypes of maternal CVD | | | | | |
| Cerebrovascular disease | 195 | 10.2 | 1.39 (1.20-1.62) | 1.18 (1.01-1.37) | 1.18 (1.02-1.37) |
| Arrhythmia | 1270 | 9.6 | 1.25 (1.17-1.32) | 1.12 (1.05-1.19) | 1.12 (1.06-1.19) |
| Heart failure | 68 | 11.3 | 1.48 (1.14-1.93) | 1.30 (0.99-1.70) | 1.30 (0.99-1.70) |
| Valvular heart disease | 299 | 8.3 | 1.15 (1.02-1.30) | 1.12 (0.99-1.26) | 1.12 (0.99-1.26) |
| Congenital heart disease | 389 | 10.8 | 1.44 (1.29-1.60) | 1.19 (1.06-1.32) | 1.19 (1.07-1.33) |
| | **ASD** | | | | |
| Composite maternal CVD | | | | | |
| No | 18 296 | 1.6 | 1.00 (Reference) | 1.00 (Reference) | 1.00 (Reference) |
| Yes | 611 | 2.7 | 1.27 (1.16-1.38) | 1.16 (1.06-1.27) | 1.17 (1.07-1.27) |
| Subtypes of maternal CVD | | | | | |
| Cerebrovascular disease | 57 | 2.8 | 1.27 (0.94-1.72) | 1.09 (0.81-1.47) | 1.09 (0.81-1.48) |
| Arrhythmia | 404 | 2.9 | 1.27 (1.14-1.41) | 1.17 (1.05-1.30) | 1.17 (1.06-1.30) |
| Heart failure | 19 | 3.0 | 1.23 (0.75-2.00) | 1.09 (0.67-1.77) | 1.09 (0.67-1.77) |
| Valvular heart disease | 73 | 1.9 | 1.21 (0.94-1.55) | 1.14 (0.89-1.46) | 1.14 (0.89-1.46) |
| Congenital heart disease | 117 | 3.0 | 1.30 (1.07-1.59) | 1.20 (0.98-1.46) | 1.20 (0.98-1.46) |
| | **ID** | | | | |
| Composite maternal CVD | | | | | |
| No | 4005 | 0.3 | 1.00 (Reference) | 1.00 (Reference) | 1.00 (Reference) |
| Yes | 96 | 0.4 | 1.16 (0.95-1.42) | 1.07 (0.88-1.31) | 1.08 (0.88-1.32) |
| Subtypes of maternal CVD | | | | | |
| Cerebrovascular disease | 13 | 0.6 | 1.77 (1.03-3.05) | 1.56 (0.91-2.67) | 1.56 (0.91-2.68) |
| Arrhythmia | 62 | 0.4 | 1.19 (0.93-1.53) | 1.11 (0.87-1.43) | 1.12 (0.87-1.44) |
| Heart failure | <5 | 0.6 | - | - | - |
| Valvular heart disease | 9 | 0.2 | 0.65 (0.34-1.24) | 0.65 (0.34-1.25) | 0.65 (0.34-1.25) |
| Congenital heart disease | 18 | 0.5 | 1.28 (0.80-2.02) | 1.08 (0.68-1.71) | 1.09 (0.69-1.73) |

Note: ADHD, attention-deficit/hyperactivity disorder; ASD, autism spectrum disorder; CI, confidence interval; CVD, cardiovascular disease; HR, hazard ratio; ID, intellectual disability.
*Incidence rates per 1000 child-years.
Model 1 was minimally adjusted for child's age, birth year and sex. Model 2 was further adjusted for mother's age at delivery, parity, region of birth, marital status, pre-gestational diabetes, pre-gestational hypertension, and parental history of any neurodevelopmental or psychiatric disorders. Model 3 was additionally adjusted for parental neighbourhood income quintiles.
-Not estimated due to insufficient events

**eTable 6. Multiple-imputation analysis of the associations between pre-existing maternal cardiovascular diseases and offspring's neurodevelopmental disorders: singleton offspring live-born without major malformations in Sweden 1990 to 2019 (N=2 940 626)**

| Exposure/s | Outcomes | | | |
|---|---|---|---|---|
| | **ADHD** | | | |
| | No. of events | Rates* | Model 1 HR (95% CI) | Model 2 HR (95% CI) |
| Composite maternal CVD | | | | |
| No | 168 725 | 4.1 | 1.00 (Reference) | 1.00 (Reference) |
| Yes | 1125 | 5.3 | 1.28 (1.21-1.36) | 1.16 (1.09-1.23) |
| Subtypes of maternal CVD | | | | |
| Cerebrovascular disease | 222 | 6.0 | 1.40 (1.22-1.59) | 1.20 (1.05-1.37) |
| Arrhythmia | 603 | 5.1 | 1.22 (1.13-1.32) | 1.14 (1.05-1.23) |
| Heart failure | 28 | 6.3 | 1.55 (1.07-2.24) | 1.39 (0.96-2.02) |
| Valvular heart disease | 93 | 4.8 | 1.15 (0.94-1.41) | 1.13 (0.92-1.38) |
| Congenital heart disease | 269 | 5.6 | 1.39 (1.23-1.57) | 1.18 (1.05-1.33) |
| | **ASD** | | | |
| Composite maternal CVD | | | | |
| No | 70 274 | 1.7 | 1.00 (Reference) | 1.00 (Reference) |
| Yes | 496 | 2.3 | 1.19 (1.09-1.30) | 1.11 (1.01-1.21) |
| Subtypes of maternal CVD | | | | |
| Cerebrovascular disease | 106 | 2.8 | 1.46 (1.20-1.76) | 1.28 (1.06-1.55) |
| Arrhythmia | 256 | 2.2 | 1.07 (0.95-1.21) | 1.01 (0.90-1.15) |
| Heart failure | 20 | 4.4 | 2.08 (1.33-3.24) | 1.90 (1.22-2.96) |
| Valvular heart disease | 49 | 2.5 | 1.16 (0.87-1.55) | 1.15 (0.86-1.53) |
| Congenital heart disease | 118 | 2.4 | 1.18 (0.98-1.42) | 1.11 (0.92-1.33) |
| | **ID** | | | |
| Composite maternal CVD | | | | |
| No | 25 315 | 0.6 | 1.00 (Reference) | 1.00 (Reference) |
| Yes | 150 | 0.7 | 1.10 (0.94-1.30) | 1.03 (0.87-1.21) |
| Subtypes of maternal CVD | | | | |
| Cerebrovascular disease | 31 | 0.8 | 1.27 (0.89-1.81) | 1.10 (0.77-1.56) |
| Arrhythmia | 66 | 0.6 | 0.84 (0.66-1.08) | 0.83 (0.65-1.06) |
| Heart failure | 5 | 1.1 | - | - |
| Valvular heart disease | 21 | 1.1 | 1.59 (1.02-2.48) | 1.41 (0.90-2.19) |
| Congenital heart disease | 41 | 0.8 | 1.23 (0.90-1.69) | 1.17 (0.85-1.60) |

Note: ADHD, attention-deficit/hyperactivity disorder; ASD, autism spectrum disorder; CI, confidence interval; CVD, cardiovascular disease; HR, hazard ratio; ID, intellectual disability.
*Incidence rates per 1000 child-years.
Model 1 was minimally adjusted for child's age, birth year and sex. Model 2 was further adjusted for mother's age at delivery, parity, education, region of birth, cohabitation with partner, smoking during early pregnancy, pre-gestational diabetes, pre-gestational hypertension, and parental history of any neurodevelopmental or psychiatric disorders.
-Not estimated due to insufficient events

**eTable 7. Sensitivity analysis restricted to later-born cohorts, concerning the associations between pre-existing maternal cardiovascular diseases and offspring's neurodevelopmental disorders: singleton offspring live-born without major malformations in Sweden 1997 to 2019 (N=2 0374 62).**

| Exposure/s | Outcomes | | | |
|---|---|---|---|---|
| | **ADHD** | | | |
| | No. of events | Rates* | Model 1 HR (95% CI) | Model 2 HR (95% CI) |
| Composite maternal CVD | | | | |
| No | 105 423 | 5.0 | 1.00 (Reference) | 1.00 (Reference) |
| Yes | 925 | 5.6 | 1.27 (1.19-1.36) | 1.15 (1.08-1.23) |
| Subtypes of maternal CVD | | | | |
| Cerebrovascular disease | 171 | 6.3 | 1.37 (1.18-1.59) | 1.18 (1.01-1.37) |
| Arrhythmia | 506 | 5.3 | 1.22 (1.11-1.33) | 1.13 (1.04-1.24) |
| Heart failure | 23 | 6.6 | 1.57 (1.04-2.36) | 1.38 (0.92-2.08) |
| Valvular heart disease | 72 | 4.7 | 1.07 (0.85-1.35) | 1.05 (0.83-1.32) |
| Congenital heart disease | 215 | 5.7 | 1.35 (1.18-1.55) | 1.16 (1.01-1.33) |
| | **ASD** | | | |
| Composite maternal CVD | | | | |
| No | 45 756 | 2.1 | 1.00 (Reference) | 1.00 (Reference) |
| Yes | 412 | 2.5 | 1.17 (1.06-1.29) | 1.09 (0.99-1.20) |
| Subtypes of maternal CVD | | | | |
| Cerebrovascular disease | 83 | 3.0 | 1.37 (1.15-1.78) | 1.27 (1.02-1.57) |
| Arrhythmia | 222 | 2.3 | 1.10 (0.96-1.25) | 1.04 (0.91-1.19) |
| Heart failure | 17 | 4.8 | 2.31 (1.44-3.72) | 2.07 (1.29-3.34) |
| Valvular heart disease | 36 | 2.3 | 1.13 (0.82-1.57) | 1.09 (0.78-1.50) |
| Congenital heart disease | 94 | 2.5 | 1.18 (0.97-1.45) | 1.10 (0.90-1.34) |
| | **ID** | | | |
| Composite maternal CVD | | | | |
| No | 13 740 | 0.6 | 1.00 (Reference) | 1.00 (Reference) |
| Yes | 114 | 0.7 | 1.05 (0.88-1.27) | 1.00 (0.83-1.20) |
| Subtypes of maternal CVD | | | | |
| Cerebrovascular disease | 20 | 0.7 | 1.13 (0.73-1.76) | 1.00 (0.65-1.55) |
| Arrhythmia | 50 | 0.5 | 0.80 (0.61-1.06) | 0.79 (0.60-1.05) |
| Heart failure | <5 | 1.1 | - | - |
| Valvular heart disease | 16 | 1.0 | 1.64 (1.01-2.68) | 1.43 (0.88-2.34) |
| Congenital heart disease | 33 | 0.9 | 1.35 (0.96-1.90) | 1.26 (0.90-1.78) |

Note: ADHD, attention-deficit/hyperactivity disorder; ASD, autism spectrum disorder; CI, confidence interval; CVD, cardiovascular disease; HR, hazard ratio; ID, intellectual disability.
*Incidence rates per 1000 child-years.
Model 1 was minimally adjusted for child's age, birth year and sex. Model 2 was further adjusted for mother's age at delivery, parity, education, region of birth, cohabitation with partner, smoking during early pregnancy, pre-gestational diabetes, pre-gestational hypertension, and parental history of any neurodevelopmental or psychiatric disorders.
-Not estimated due to insufficient events

**eTable 8. Sensitivity analysis restricted to later-born cohorts, concerning the associations between pre-existing maternal cardiovascular diseases and offspring's neurodevelopmental disorders: singleton offspring live-born without major malformations in British Columbia, Canada 2001 to 2019 (N= 589 933).**

| **Exposure/s** | **Outcomes** | | | |
|---|---|---|---|---|
| | **ADHD** | | | |
| | No. of events | Rates* | Model 1<br>HR (95% CI) | Model 2<br>HR (95% CI) |
| Composite maternal CVD | | | | |
| No | 40 389 | 8.2 | 1.00 (Reference) | 1.00 (Reference) |
| Yes | 1430 | 10.5 | 1.31 (1.24-1.39) | 1.14 (1.08-1.21) |
| Subtypes of maternal CVD | | | | |
| Cerebrovascular disease | 136 | 11.0 | 1.40 (1.17-1.68) | 1.18 (0.99-1.42) |
| Arrhythmia | 926 | 10.2 | 1.26 (1.18-1.35) | 1.12 (1.05-1.20) |
| Heart failure | 54 | 12.1 | 1.57 (1.18-2.09) | 1.31 (0.98-1.77) |
| Valvular heart disease | 178 | 11.3 | 1.35 (1.16-1.58) | 1.27 (1.08-1.48) |
| Congenital heart disease | 280 | 11.4 | 1.44 (1.26-1.64) | 1.13 (0.99-1.28) |
| | **ASD** | | | |
| Composite maternal CVD | | | | |
| No | 13 596 | 2.7 | 1.00 (Reference) | 1.00 (Reference) |
| Yes | 527 | 3.7 | 1.29 (1.17-1.41) | 1.19 (1.08-1.31) |
| Subtypes of maternal CVD | | | | |
| Cerebrovascular disease | 54 | 4.2 | 1.41 (1.03-1.92) | 1.22 (0.89-1.66) |
| Arrhythmia | 351 | 3.7 | 1.27 (1.14-1.42) | 1.18 (1.06-1.32) |
| Heart failure | 17 | 3.6 | 1.23 (0.73-2.08) | 1.09 (0.64-1.84) |
| Valvular heart disease | 55 | 3.3 | 1.29 (0.96-1.73) | 1.20 (0.89-1.61) |
| Congenital heart disease | 102 | 4.0 | 1.30 (1.05-1.61) | 1.19 (0.96-1.48) |
| | **ID** | | | |
| Composite maternal CVD | | | | |
| No | 1959 | 0.4 | 1.00 (Reference) | 1.00 (Reference) |
| Yes | 70 | 0.5 | 1.31 (1.03-1.66) | 1.21 (0.95-1.53) |
| Subtypes of maternal CVD | | | | |
| Cerebrovascular disease | 11 | 0.8 | 2.29 (1.27-4.14) | 2.00 (1.11-3.59) |
| Arrhythmia | 47 | 0.5 | 1.31 (0.98-1.75) | 1.25 (0.94-1.68) |
| Heart failure | <5 | - | - | - |
| Valvular heart disease | 5 | 0.3 | 0.76 (0.32-1.83) | 0.76 (0.31-1.82) |
| Congenital heart disease | 12 | 0.5 | 1.25 (0.71-2.21) | 1.02 (0.57-1.79) |

Note: ADHD, attention-deficit/hyperactivity disorder; ASD, autism spectrum disorder; CI, confidence interval; CVD, cardiovascular disease; HR, hazard ratio; ID, intellectual disability.
*Incidence rates per 1000 child-years.
Model 1 was minimally adjusted for child's age, birth year and sex. Model 2 was further adjusted for mother's age at delivery, parity, region of birth, marital status, pre-gestational diabetes, pre-gestational hypertension, and parental history of any neurodevelopmental or psychiatric disorders.
-Not estimated due to insufficient events

**eTable 9. Incidence rates and hazard ratios of autism spectrum disorder without intellectual disability according to pre-existing maternal cardiovascular diseases: singleton offspring live-born without major malformations in Sweden 1990 to 2019 (N=2 672 229) and British Columbia, Canada 1992 to 2019 (N= 887 582).**

| | **Sweden** | | | |
|---|---|---|---|---|
| **Exposure/s** | No. of events | Rates* | Model 1 HR (95% CI) | Model 2 HR (95% CI) |
| Composite maternal CVD | | | | |
| No | 54 907 | 1.4 | 1.00 (Reference) | 1.00 (Reference) |
| Yes | 402 | 2.0 | 1.23 (1.11-1.36) | 1.14 (1.03-1.25) |
| Subtypes of maternal CVD | | | | |
| Cerebrovascular disease | 86 | 2.5 | 1.53 (1.24-1.89) | 1.34 (1.09-1.66) |
| Arrhythmia | 215 | 1.9 | 1.16 (1.01-1.33) | 1.08 (0.95-1.24) |
| Heart failure | 14 | 3.4 | 2.06 (1.22-3.48) | 1.90 (1.12-3.20) |
| Valvular heart disease | 32 | 1.8 | 1.10 (0.78-1.55) | 1.06 (0.75-1.50) |
| Congenital heart disease | 92 | 2.1 | 1.26 (1.02-1.54) | 1.16 (0.94-1.42) |
| | **British Columbia, Canada** | | | |
| Composite maternal CVD | | | | |
| No | 16 965 | 1.5 | 1.00 (Reference) | 1.00 (Reference) |
| Yes | 574 | 2.5 | 1.27 (1.17-1.39) | 1.16 (1.06-1.27) |
| Subtypes of maternal CVD | | | | |
| Cerebrovascular disease | 52 | 2.5 | 1.25 (0.92-1.70) | 1.06 (0.78-1.45) |
| Arrhythmia | 380 | 2.7 | 1.28 (1.15-1.42) | 1.17 (1.05-1.30) |
| Heart failure | 17 | 2.6 | 1.15 (0.70-1.90) | 1.02 (0.62-1.67) |
| Valvular heart disease | 70 | 1.8 | 1.27 (0.98-1.64) | 1.18 (0.92-1.53) |
| Congenital heart disease | 112 | 2.8 | 1.32 (1.08-1.62) | 1.21 (0.99-1.49) |

Note: CI, confidence interval; CVD, cardiovascular disease; HR, hazard ratio.
*Incidence rates per 1000 child-years.
Model 1 was minimally adjusted for child’s age, birth year and sex. Model 2 was further adjusted for mother’s age at delivery, parity, education (Sweden only), region of birth, marital status/cohabitation with partner, smoking during early pregnancy (Sweden only), pre-gestational diabetes, pre-gestational hypertension, and parental history of any neurodevelopmental or psychiatric disorders.
